# Oral colonization by *Entamoeba gingivalis* and *Trichomonas* tenax: PCR-based study in health, gingivitis, and periodontitis

**DOI:** 10.1101/2020.07.16.20155861

**Authors:** Alaa’ Yaseen, Azmi Mahafzah, Deema Dababseh, Duaa Tayem, Ahmad A. Hamdan, Esraa Al-Fraihat, Yazan Hassona, Gülşen Özkaya Şahin, Julien Santi-Rocca, Malik Sallam

## Abstract

The etiology of periodontitis needs further investigation, as is the place of gingivitis in its pathophysiology. A few studies linked the oral colonization by parasites (*Entamoeba gingivalis* and *Trichomonas tenax*) to the disease and its severity. The aim of this study was to estimate the prevalence of these oral parasites among healthy individuals, gingivitis and periodontitis patients in Jordan. The study was conducted by active enrolment of participants at Jordan University Hospital. The participants answered a questionnaire that included items related to possible risk factors for periodontal disease. Saliva and dental plaque samples were collected. The detection of oral parasites was done using conventional PCR and microscopic examination of wet mounts. The study population comprised a total of 237 individuals divided into three groups: healthy (n=94), gingivitis (n=53), and periodontitis (n=90). PCR results revealed that the overall prevalence of *E. gingivalis* was 71.7% compared to 12.2% for *T. tenax*. The periodontal disease group had higher prevalence of *E. gingivalis* and *T. tenax* compared to the healthy group (p<0.001). Increasing age was associated with higher prevalence of *E. gingivalis* (p=0.008) and *T. tenax* (p=0.019), in the entire study population. The number of cases of colonization detected by microscopic observation was lower for any of the oral parasites, as compared to diagnosis by PCR (40.7% vs. 71.7%, p<0.001 for *E. gingivalis* and 4.3% vs. 12.2%, p=0.007 for *T. tenax*). The higher prevalence of oral parasites among patients with periodontal disease might point to their potential contribution in the disease and its severity.

**IMPORTANCE:** Periodontal disease has a high prevalence globally, with adverse effects on the quality of life for affected individuals. Despite the presence of several studies that investigated the role of oral parasites in periodontal disease, reliable conclusions about this matter remained elusive mainly due to utilization of microscopy in parasite detection. The current study provides new insights into the epidemiology and prevalence of the two oral parasites (*Entamoeba gingivalis* and *Trichomonas tenax*) in patients with various stages of periodontal disease in comparison to healthy adults. In addition, we describe the potential role of oral colonization by parasites as a risk factor for development of periodontal disease and its severity using a molecular-based approach.

## Introduction

In recent years, the role of microbiota in establishing and maintaining a healthy status among humans has been demonstrated by molecular methods (1-3). The oral microbiota was no exception, and its disruption has been shown to result in a various range of oral diseases including gingivitis and periodontitis (4, 5). This disruption in oral microbiota is termed dysbiosis, in contrast to eubiosis defined as the complex interactions of different resident microbes that result in an equilibrium to maintain the healthy state of the ecologic niche, which is the oral cavity in this case (4-7).

Periodontal disease represents a state of chronic inflammation in gingiva, bone and supporting ligaments (8, 9). The physiologic healthy state of gingiva can be defined as the total absence or minimal levels of clinical inflammation of the periodontium with normal support (no loss affecting attachment or bone) (10, 11). The identification of plaque-induced gingivitis relies on the presence of bleeding on probing with an intact periodontium and/or visible inflammation and this condition can be reversed back to a healthy state if managed properly (11, 12). Periodontitis results in destruction of the periodontal ligament, cementum and alveolar bone. Inflammation and microbiota of periodontitis can be cured but tissues are not healed back to their initial volume, organization, and shape, thus necessitating continuous maintenance of good oral hygiene (11, 13-15).

Despite the lack of accurate and recent estimates of its prevalence, periodontal disease is considered among the most common conditions affecting all age groups with predilection for the elderly (6, 16). As of 2010, the prevalence of periodontitis was 47% among adults aged 30 and above in the United States, while the global prevalence of severe periodontitis was 11%, with higher estimates for gingivitis (17-19). In addition, periodontitis is considered an important cause of teeth loss in older adults, which adversely affects the quality of life (20). The underlying etiology and pathogenesis of periodontal disease has been linked to microbial dysbiosis (21, 22). However, the exact specific roles of different microbes in the dental plaque that lead to the development of periodontal disease remains an enigma (7, 23). In addition, the initiating factors for microbial dysbiosis in the oral cavity remains unclear and the study of these factors is a subject for ongoing research (5).

Several factors have been linked to an increased incidence of periodontal disease and these can be divided into non-modifiable and modifiable factors (24). Examples of the former include aging, genetic predisposition, and osteoporosis, whereas the later include smoking, diabetes mellitus, psychological stress, alcohol consumption and poor oral hygiene (24-31).

The role of the parasitic fraction of the oral microbiome, namely: *Entamoeba gingivalis* and *Trichomonas tenax*, is considered among the proposed models regarding the contributing factors to the development of periodontal disease (32, 33). Several studies aimed to investigate parasitic oral colonization among healthy individuals and those with periodontal disease with remarkably variable results (34-41). Such variability can be related to adoption of different approaches for parasite detection, the existence of previously unknown genetic variants of oral parasites, in addition to limitations of small sample sizes, and possible bias in selection of study subjects among others as reviewed recently (42, 43).

Thus, the objective of the current study was to examine potential differences of the two reference methods for oral parasite detection (microscopy vs. PCR), which might help to explain the differences observed between studies. In addition, we aimed to investigate the prevalence of both parasites in health and periodontal disease, to check for possible regional specificities in Jordan. Also, we aimed to better define the place of gingivitis in the physiopathology of periodontal disease using parasite colonization. Finally, we aimed to identify the variables that might be associated with increased likelihood of harboring these parasites in health and disease.

## Results

### Study population

The total number of study participants who were eligible to be included in the final analysis was 237, distributed as follows: healthy group (n=94, 39.7%), gingivitis group (n=53, 22.4%) and periodontitis group (n=90, 38.0%, Table 1). Significant differences among the three study groups were found for the following factors: the median age of the healthy group was younger compared to the two other groups combined (24 vs. 44 years, p<0.001, Mann-Whitney U test [M-W]). The periodontitis group had an older median age compared to the two other groups (p<0.001, Kruskall-Wallis test [K-W], Table 1). The percentage of smokers in the healthy group was lower in comparison to the disease group with ex-smokers excluded (31.9% vs. 50.3%, p=0.004, χ^2^ test). The monthly income was the lowest among the periodontitis group and the highest among the healthy group (p<0.001, K-W). In addition, the periodontitis group had the lowest overall level of dental care (p<0.001, K-W) and the highest body mass index (BMI) levels (p=0.003, K-W, Table 1).

**Table 1.**
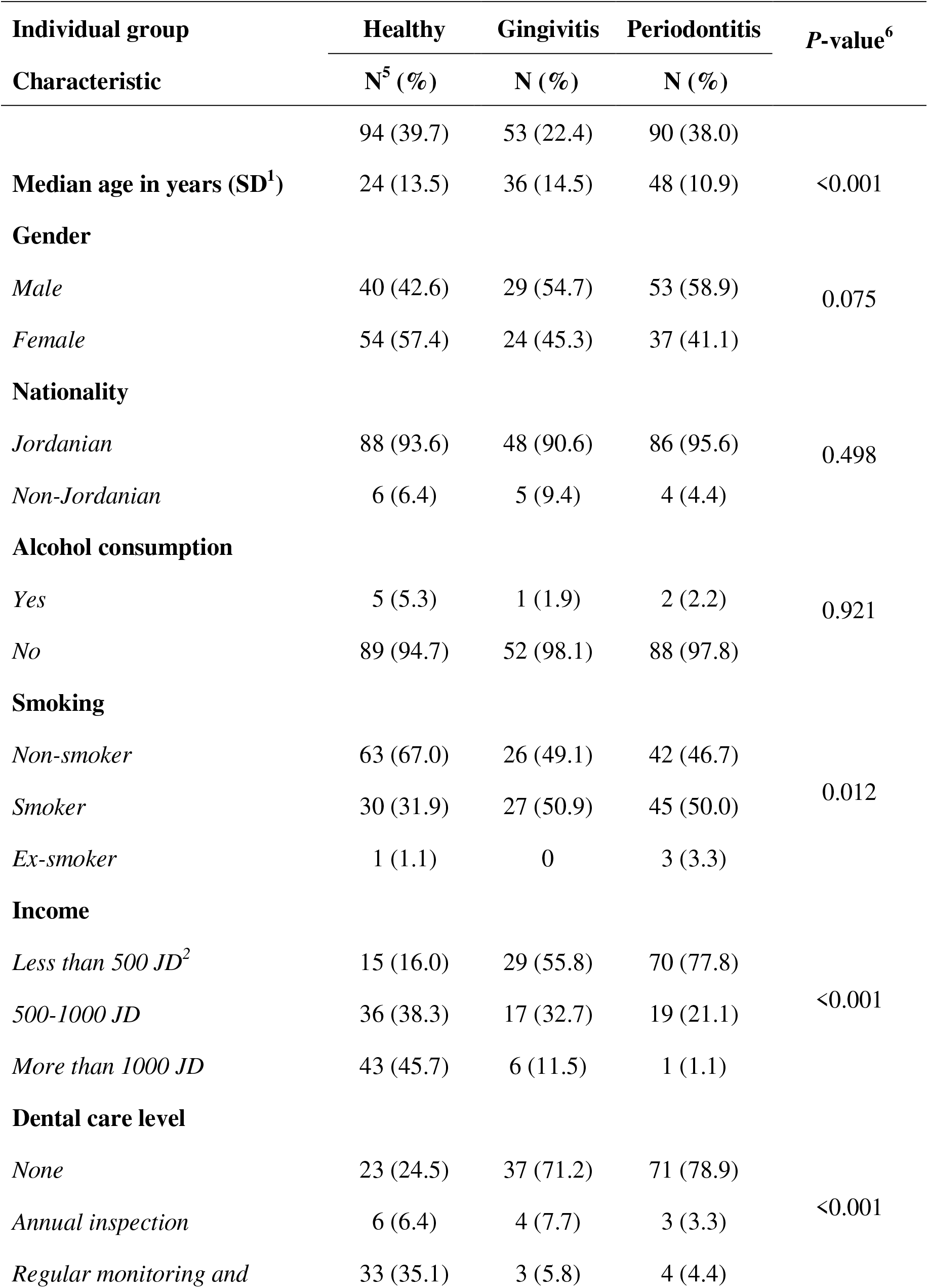
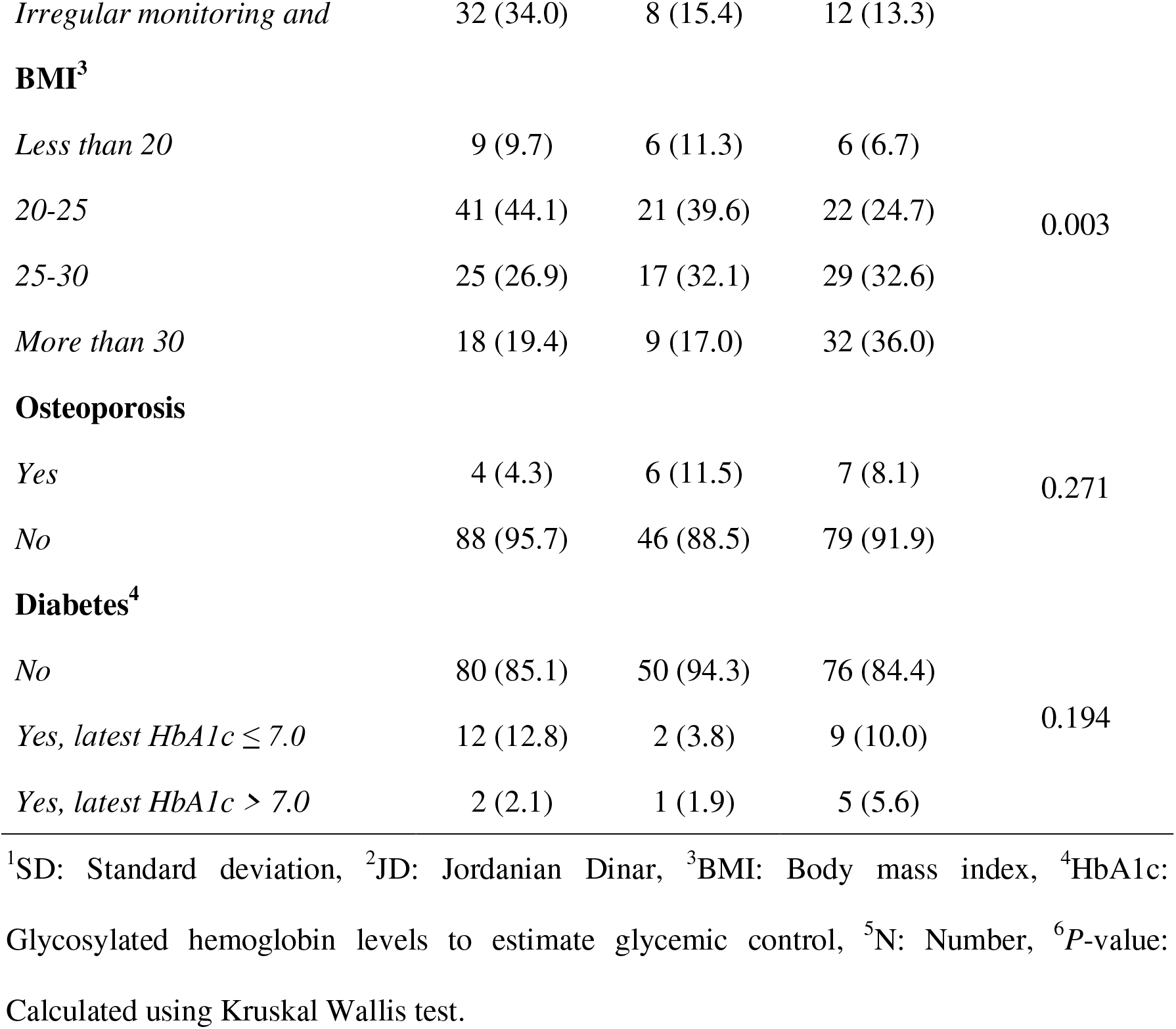
Characteristics of the study population divided by the three study groups.

### The prevalence of oral parasites in the study population

To estimate the overall prevalence of *E. gingivalis* and *T. tenax*, the conventional PCR results were available from the 237 individuals of the three groups for final analysis. The overall prevalence of *E. gingivalis* among the entire study population was 71.7% (95% confidence interval [CI]: 65.7% to 77.1%), while the overall prevalence for *T. tenax* was 12.2% (95% CI: 8.6% to 17.1%).

Concurrent detection of the two oral parasites (dual colonization) was detected in 26 study subjects yielding a prevalence of 11.0% (95% CI: 7.6% to 15.6%). Among the 29 study subjects with *T. tenax* colonization, *E. gingivalis* was also present in 26 individuals (89.7%). Divided by the study groups, the prevalence of *E. gingivalis* was the highest among the periodontitis group (n=80, 88.9%) compared to gingivitis (n=45, 84.9%) and healthy group (n=45, 47.9%). The difference was statistically significant upon comparing the healthy group to gingivitis and periodontitis groups (p<0.001 for both comparisons, χ^2^ test). However, the difference was not significant upon comparing the gingivitis group with the periodontitis group (p=0.603, χ^2^ test). For *T. tenax*, the prevalence increased starting from 3.2% in the heathy group, to 5.7% in the gingivitis group and ending in 25.6% in the periodontitis group. The difference was not significant by comparing the healthy and gingivitis groups (p=0.668, χ^2^ test), but significant results were noticed upon comparing the periodontitis group to healthy and gingivitis groups (p<0.001 and 0.003 respectively for the two comparisons, χ^2^ test).

### Factors associated with higher prevalence of oral parasites in the whole study population

The entire study population comprised 122 males and 115 females. *T. tenax* was detected more frequently among males compared to females (16.4% vs. 7.8%, p=0.044, χ^2^ test). In addition, *E. gingivalis* and dual colonization showed the following gender prevalence, that lacked statistical significance (74.6% vs. 68.7%; 14.8% vs. 7.0%, p=0.314; p=0.055, χ^2^ test).

For both oral parasites *E. gingivalis* and *T. tenax*, higher prevalence of colonization was found among the study subjects with the lowest income (p<0.001 for *E. gingivalis*, p=0.010 for *T. tenax*, χ^2^ test). No previous history of dental care was associated with higher prevalence of *E. gingivalis* (p<0.001, χ^2^ test), whereas higher levels of colonization by *T. tenax* was associated with Non-Jordanian nationality (p=0.010, χ^2^ test) and higher BMI (p=0.012, χ^2^ test).

The bleeding index levels were higher among study subjects positive for *E. gingivalis* and *T. tenax* (mean: 0.53 vs. 0.20, and 0.73 vs. 0.39, respectively p<0.001, M-W). For the plaque index, significant difference was only found among individuals positive for *E. gingivalis* (mean: 0.79 vs. 0.55, p<0.001, M-W). The following factors did not give significant differences in colonization by either of the two oral parasites: smoking, smoking exposure through pack-year evaluation, history of diabetes, family history of gum disease, alcohol use and osteoporosis.

### Factors associated with higher prevalence of oral parasites in each study group

The analysis of the variables associated with higher prevalence of colonization by oral parasites divided by the three groups revealed the following significant results: In the healthy group, *E. gingivalis* was detected in 100.0% of the individuals with uncontrolled diabetes compared to 75.0% among those with controlled diabetes and 42.5% among those without history of diabetes (p=0.012, χ^2^ test). In the periodontitis group, all study subjects with non-Jordanian nationality were positive for *T. tenax* (n=4) compared to 22.1% among Jordanians (p<0.001, χ^2^ test) and individuals with BMIs higher than 25 had a prevalence of *E. gingivalis* of 93.4% compared to 78.6% among individuals with BMIs less than 25 (p=0.039, χ^2^ test) and a similar pattern was observed for *T. tenax* with 10.7% among individuals with BMIs less than 25 vs. 32.8% among individuals with BMIs higher than 25 (p=0.027, χ^2^ test).

### Association of oral parasites with periodontal disease severity

Data on the type of periodontal disease (localized vs. generalized) was available from 130 study subjects. The localized type comprised 18 individuals as opposed to 112 individuals with generalized periodontal disease. *T. tenax* was only detected among individuals with generalized periodontal disease compared to its total absence among those with localized periodontal disease (19.6% vs 0.0%. p=0.039, χ^2^ test) and no dual colonization was detected either in the localized group compared to generalized group (p=0.051, χ^2^ test). Despite the higher prevalence of both oral parasites in individuals with advanced stage and grade of disease, no statistical significance was detected.

### Comparing the accuracy of microscopic examination to molecular detection for oral parasites

To assess the sensitivity of microscopic examination of wet mounts in the evaluation of colonization by oral parasites to molecular detection using conventional PCR, we compared the rate of positivity for each method. Microscopic examination detected *E. gingivalis* in 94 out of 231 (40.7%) specimens analyzed using this method compared to positivity rate of 71.7% using PCR (p<0.001, χ^2^ test) and missing a total of 76 cases. For *T. tenax*, microscopic examination detected only 10 out of 231 (4.3%) specimens compared to 12.2% using PCR (p=0.002, χ^2^ test) and missing a total of 19 cases. Stratified by the study group, the largest proportion of missed cases of *E. gingivalis* was noticed among healthy group (73.3%) compared to gingivitis group (40.5%) and periodontitis group (26.0%, p<0.001, linear-by-linear test for association [LBL]). For *T. tenax*, all cases were missed in the healthy and gingivitis groups, while the missing rate was 56.5% in the periodontitis group (p=0.066, LBL).

### Risk factors for periodontal disease in the study population

The majority of risk factors for periodontal disease that were previously reported in various studies were tested in this work (e.g. dental care level, smoking, DM, etc.). To analyse the patterns associated with higher likelihood of having periodontal disease as a whole and per disease state (gingivitis and periodontitis), we conducted multinomial logistic regression analysis using variables that were classified into dichotomous outcomes as follows: For age (more than 38 years vs. less than or equal to 38 years, [38 years was the median age for the whole population]), gender (male vs. female), nationality (Jordanian vs. non-Jordanian), BMI (more than or equal to 25 vs. less than 25), monthly income (less than 500 JD vs. more than or equal to 500 JD), dental care (no dental care vs. any form of dental care), smoking (smoker vs. non-smoker, [ex-smokers were excluded]), DM (controlled or non-controlled DM vs. non-diabetic), family history (present vs. absent), subjective evaluation of stress (more stressed if the score is 0-4 vs. less stressed if the score is more than 5), alcohol use (consumer vs. non-consumer, [ex-consumers were excluded]), osteoporosis (present vs. absent), *E. gingivalis* by PCR (positive vs. negative) and *T. tenax* by PCR (positive vs. negative).

For the health vs. disease groups, the most significant risk factors for development of periodontal disease in the current study was colonization by *E. gingivalis* (p<0.001, odds ratio [OR]=5.8). Other significant risk factors included monthly income that is less than 500 JD, lack of dental care and higher levels of stress evaluated using the questionnaire (Tables 2, 3 and 4). In gingivitis compared to healthy individuals, significant risk factors included lack of dental care, colonization by *E. gingivalis* and low monthly income, while for periodontitis vs. healthy individuals, colonization by each oral parasite was an independent risk factor for the disease (p=0.007, OR=5.2 for *E. gingivalis* and p=0.009, OR=10.1 for *T. tenax*, Tables 2, 3 and 4).

**Table 2.** Potential risk factors for periodontal disease upon comparing the healthy individuals vs. those with periodontal disease (gingivitis or periodontitis)

| **Potential risk factors for periodontal disease vs. healthy individuals** | ***P* value<sup>9</sup>** | **OR<sup>10</sup>** | **95% CI<sup>11</sup>** |
| --- | --- | --- | --- |
| ***Entamoeba gingivalis* by PCR (positive vs. negative)<sup>1</sup>** | **<0.001** | **5.84** | **2.18-15.66** |
| **Monthly income (less than 500 JD vs. more than or equal to 500 JD)<sup>2</sup>** | **<0.001** | **5.43** | **2.18-13.50** |
| **Dental care (no dental care vs. any form of dental care)** | **0.001** | **4.45** | **1.89-10.50** |
| **Subjective evaluation of stress (more stressed vs. less stressed)<sup>3</sup>** | **0.006** | **3.87** | **1.47-10.20** |
| *Trichomonas tenax* by PCR (positive vs. negative) | 0.077 | 4.05 | 0.86-19.07 |
| Age (more than 38 years vs. less than or equal to 38 years)<sup>4</sup> | 0.180 | 2.01 | 0.73-5.54 |
| Osteoporosis (present vs. absent) | 0.466 | 1.90 | 0.34-10.63 |
| Smoking (smoker vs. non-smoker)<sup>5</sup> | 0.181 | 1.82 | 0.76-4.38 |
| Gender (male vs. female) | 0.461 | 1.40 | 0.57-3.47 |
| Family history (present vs. absent) | 0.656 | 1.25 | 0.48-3.26 |
| Nationality (Jordanian vs. Non-Jordanian) | 0.904 | 1.13 | 0.17-7.67 |
| BMI (more than or equal to 25 vs. less than 25)<sup>6</sup> | 0.754 | 0.86 | 0.35-2.16 |
| Alcohol use (consumer vs. non-consumer)<sup>7</sup> | 0.395 | 0.35 | 0.03-3.87 |
| DM (controlled or non-controlled DM vs. non-diabetic)<sup>8</sup> | 0.053 | 0.26 | 0.07-1.02 |

<sup>1</sup>PCR: Conventional polymerase chain reaction, <sup>2</sup>JD: Jordanian Dinar, <sup>3</sup>Subjective evaluation of stress: using a nine-item questionnaire, <sup>4</sup>Age: The median age for the entire study population was 38 years, <sup>5</sup>Smoking: Ex-smokers were excluded from analysis (n=4), <sup>6</sup>BMI: Body mass index, <sup>7</sup>Alcohol use: Ex-users of alcohol were excluded from analysis (n=3), <sup>8</sup>DM: Diabetes mellitus. Significant results are shown in bold, <sup>9</sup>P-value: Calculated using multinomial logistic regression and likelihood ratio tests, <sup>10</sup>OR: Odds ratio, <sup>11</sup>CI: Confidence interval.

**Table 3.** Risk factors for gingivitis and compared to healthy individuals in the current study population.

| Potential risk factors for gingivitis vs. healthy individuals | *P* value<sup>9</sup> | OR<sup>10</sup> | 95% CI<sup>11</sup> |
| --- | --- | --- | --- |
| Dental care (no dental care vs. any form of dental care) | 0.001 | 5.78 | 2.13-15.70 |
| *Entamoeba gingivalis* by PCR (positive vs. negative)<sup>1</sup> | 0.002 | 5.79 | 1.86-18.02 |
| Monthly income (less than 500 JD vs. more than or equal to 500 JD)<sup>2</sup> | 0.010 | 3.87 | 1.39-10.84 |
| DM (controlled or non-controlled DM vs. non-diabetic)<sup>3</sup> | 0.078 | 0.21 | 0.04-1.19 |
| Subjective evaluation of stress (more stressed vs. less stressed)<sup>4</sup> | 0.115 | 2.38 | 0.81-6.97 |
| Osteoporosis (present vs. absent) | 0.129 | 4.17 | 0.66-26.36 |
| Smoking (smoker vs. non-smoker)<sup>5</sup> | 0.311 | 1.66 | 0.62-4.40 |
| Gender (male vs. female) | 0.425 | 1.51 | 0.55-4.11 |
| Alcohol use (consumer vs. non-consumer)<sup>6</sup> | 0.452 | 0.38 | 0.03-4.69 |
| BMI (more than or equal to 25 vs. less than 25)<sup>7</sup> | 0.528 | 0.73 | 0.27-1.97 |
| Family history (present vs. absent) | 0.744 | 0.83 | 0.27-2.55 |
| Age (more than 38 years vs. less than or equal to 38 years)<sup>8</sup> | 0.798 | 0.86 | 0.27-2.76 |
| *Trichomonas tenax* by PCR (positive vs. negative) | 0.903 | 0.88 | 0.11-7.11 |
| Nationality (Jordanian vs. Non-Jordanian) | 0.934 | 1.09 | 0.16-7.60 |

<sup>1</sup>PCR: Conventional polymerase chain reaction, <sup>2</sup>JD: Jordanian Dinar, <sup>3</sup>DM: Diabetes mellitus, <sup>4</sup>Subjective evaluation of stress: using a nine-item questionnaire, <sup>5</sup>Smoking: Ex-smokers were excluded from analysis (n=4), <sup>6</sup>Alcohol use: Ex-users of alcohol were excluded from analysis (n=3), <sup>7</sup>BMI: Body mass index, <sup>8</sup>Age: The median age for the entire study population was 38 years. Significant results are shown in bold, <sup>9</sup>P-value: Calculated using multinomial logistic regression and likelihood ratio tests, <sup>10</sup>OR: Odds ratio, <sup>11</sup>CI: Confidence interval.

**Table 4.** Risk factors for periodontitis and compared to healthy individuals in the current study population.

| **Potential risk factors for periodontitis vs. healthy individuals** | ***P* value<sup>1</sup>** | **OR<sup>2</sup>** | **95% CI<sup>3</sup>** |
| --- | --- | --- | --- |
| **Monthly income (less than 500 JD vs. more than or equal to 500 JD)<sup>4</sup>** | **<0.001** | **7.70** | **2.65-22.33** |
| **Subjective evaluation of stress (more stressed vs. less stressed)<sup>5</sup>** | **0.001** | **6.36** | **2.12-19.14** |
| ***Entamoeba gingivalis* by PCR (positive vs. negative)<sup>6</sup>** | **0.007** | **5.21** | **1.58-17.25** |
| ***Trichomonas tenax* by PCR (positive vs. negative)** | **0.009** | **10.09** | **1.77-57.53** |
| **Age (more than 38 years vs. less than or equal to 38 years)<sup>7</sup>** | **0.011** | **4.43** | **1.40-14.05** |
| **Dental care (no dental care vs. any form of dental care)** | **0.014** | **3.68** | **1.30-10.40** |
| DM (controlled or non-controlled DM vs. non-diabetic)<sup>8</sup> | 0.102 | 0.28 | 0.06-1.29 |
| Smoking (smoker vs. non-smoker)<sup>9</sup> | 0.162 | 2.09 | 0.74-5.87 |
| Family history (present vs. absent) | 0.375 | 1.65 | 0.54-5.01 |
| Alcohol use (consumer vs. non-consumer)<sup>10</sup> | 0.416 | 0.17 | 0.01-12.59 |
| Gender (male vs. female) | 0.550 | 1.39 | 0.48-4.03 |
| Nationality (Jordanian vs. Non-Jordanian) | 0.718 | 1.56 | 0.14-17.30 |
| BMI (more than or equal to 25 vs. less than 25)<sup>11</sup> | 0.917 | 1.06 | 0.37-3.06 |
| Osteoporosis (present vs. absent) | 0.973 | 0.97 | 0.13-7.07 |

<sup>1</sup>P-value: Calculated using multinomial logistic regression and likelihood ratio tests, <sup>2</sup>OR: Odds ratio, <sup>3</sup>CI: Confidence interval, <sup>4</sup>JD: Jordanian Dinar, <sup>5</sup>Subjective evaluation of stress: using a nine-item questionnaire, <sup>6</sup>PCR: Conventional polymerase chain reaction, <sup>7</sup>Age: The median age for the entire study population was 38 years, <sup>8</sup>DM: Diabetes mellitus, <sup>9</sup>Smoking: Ex-smokers were excluded from analysis (n=4), <sup>10</sup>Alcohol use: Ex-users of alcohol were excluded from analysis (n=3), <sup>11</sup>BMI: Body mass index. Significant results are shown in bold.

## Discussion

Gingivitis and periodontitis are inflammatory conditions that can also be viewed as infectious diseases (44). In the oral microbiome, the role of the bacterial fraction in periodontitis has been studied extensively with accumulating evidence pointing to its contribution to the etiology of the disease (45). However, the minority fraction of protozoa was not studied to a similar level compared to its bacterial counterpart (32, 43, 46, 47). Thus, more studies are warranted to evaluate the potential role of the human oral “protozoome” in health and disease.

In the current study, we investigated the prevalence and risk factors for oral colonization by the currently known human oral parasites, *E. gingivalis* and *T. tenax*, for the first time in Jordan. The importance of this work is related to the following aspects: First, periodontal disease (gingivitis and periodontitis) has a high prevalence among individuals of different age groups, which poses significant risks to public health, including various infections and potential tooth loss (6, 16, 20). Second, some key elements regarding the etiology and pathophysiology of periodontal disease have not been disentangled yet, hence more research is needed to decipher these unresolved elements (5, 7, 23). Third, despite the uncertainties regarding the specific roles of different microorganisms in periodontal disease, the accumulating evidence points to conspicuous differences in the oral microbiome between health and disease and the role of oral parasites in both states is yet to be clearly delineated (43). Fourth, a few studies on the epidemiology of oral parasites originated from the Middle East and North Africa (MENA) region, including studies from Egypt, Iran, Iraq and Saudi Arabia, without relying on molecular detection for estimating the burden of these oral parasites in the countries of the region (35, 37-39, 48, 49). Thus, we aimed to build on the previous work that had the same objective, while attempting to avoid some limitations of the previously published reports. For example, in this study, each sample was based on a mixture of saliva and dental plaques as opposed to relying on one of them solely for parasite detection. The advantage of this sampling approach is related to improving the sensitivity of detection as oral parasites can be found in either one of these sites (50). Also, we relied on contemporaneous use of the two reference methods currently applied for oral parasite detection (34, 43). In addition, we tried to improve the sensitivity of molecular detection in this study through using silica column-based DNA extraction method, which can help to remove PCR inhibitors and we adopted sensitive PCR protocols that were previously validated (34, 51, 52).

The main result of the study was the finding of a profoundly higher prevalence of *E. gingivalis* (87.4%) among individuals with periodontal disease, compared to those in the healthy group (47.9%) using the PCR method. For *T. tenax*, the estimates were much lower and significant differences were found moving from 3.2% in the healthy group to 18.2% among individuals with periodontal disease. To assess reproducibility of these results, a limited number of studies were found (34, 36, 43, 53, 54). The comparisons were further complicated by the reliance of a majority of the previous studies on microscopic detection methods (examination of wet mounts or permanent stained smears) (35, 37-41). In the current study, a significant number of colonization cases by the two oral parasites was missed upon using the microscopic approach. One possible explanation for this discrepancy is the subjectivity of the microscopic approach which depends on the skills and experience of the examiner, number of the fields examined, method of microscopy used (light vs. phase-contrast), use of staining, nature of mounting media, and the lag time between sampling and examination (particularly for wet mount examination which depends on the viability of oral parasites, since motility is one of the decisive defining features for diagnosis) (34).

Despite the variability in results of the previously published reports, two recurring patterns were observed and were in line with our results. First, the observation of an increase in the prevalence of both oral parasites moving from health to gingivitis and reaching the highest levels in periodontitis (43). Second, the generally higher prevalence of *E. gingivalis* in comparison to *T. tenax* in both health and disease. Interestingly, a significant association between the presence of *T. tenax* and periodontal disease severity was also found which was manifested in its total absence in localized disease. This result should be interpreted with extreme caution taking into account the limited number of individuals with localized disease that were included in the study (n=12). However, Marty *et al* hinted to the potential existence of an association between oral colonization by *T. tenax* and severity of periodontal disease, thus, our observation might not appear as an unforeseen result (33, 47). Since the current consensus is the belief that the role of microbial communities rather than single microbes are implicated in the development of periodontal disease, it appears that the significant differences observed in this study among different groups and for the two oral parasites is genuine and the potential pathogenic roles of these oral parasites should be dissected continuously similar to recent work by Bao *et al* (21, 55).

In the few studies that used the same molecular approach (conventional PCR), for identification of oral parasites, a couple reported a prevalence of *E. gingivalis* that was consistent with our results (34, 54, 56). Bonner *et al* reported a slightly lower prevalence of 33.3% for *E. gingivalis* among healthy individuals whereas Garcia *et al* results were close at 48.6%. In the two other remaining studies, *E. gingivalis* was not detected at all among healthy individuals, however, these two studies suffered from two shortcomings. First, they used different sets of primers that might resulted in missing some cases particularly those with possible low parasite loads. Second, the two studies included smaller sample sizes (12 and 20 samples) (36, 53). For periodontitis, results were available from three studies only, and the prevalence of *E. gingivalis* ranged from 26.9% to 80.6% (34, 36, 54). Three studies that used PCR for the detection of *T. tenax*, reported higher prevalence among individuals with periodontitis (39, 47, 57, 58). However, the reported prevalence in these studies varied considerably (26.9% vs. 40.0% and 70.0%), which might be related to the differences in the study populations. Another explanation might be the presence of regional specificities in this type of colonization since we found a statistically significant higher prevalence of *T. tenax* among the study subjects of non-Jordanian countries of origin (all where from other MENA countries).

Analysis of different individual variables for possible association with increased likelihood of harbouring the oral parasites was futile to say the least. Patterns in the whole population differed when analysis was done by stratification into the three individual groups, and also no specific patterns were consistently found in other studies (37, 58, 59). However, an interesting observation that can be seen in this study is that colonization by oral parasites *per se* appeared to be an independent risk factor for periodontal disease. Indirect indicators of a lower socio-economic status (low income and absence of previous dental care) appeared to have the most obvious association with higher prevalence of oral parasites besides the increasing age irrespective of the individual group (which might be related to an increased likelihood of exposure). The known risk factors for periodontitis were not necessarily associated with higher prevalence of oral parasites, which makes us inclined to propose that the presence of oral parasites may not merely be a marker of the disease and might rather play a larger role that has not been appreciated yet.

Limitations of the current work were inevitable and included difficulty in matching different groups (health and disease), particularly for age and income levels, which precluded conducting the study in a case-control type. The application of quantitative PCR could have resolved association between the parasite load and periodontal disease, especially for *E. gingivalis*, and this should be considered in any future work trying to link oral parasites in health and disease, especially with availability of an experimentally validated protocol for such an aim (60). Also, the strain variability particularly for *E. gingivalis* was not covered completely in this work since we did not use the ST2 primers aimed at the detection of the second currently known variant of *E. gingivalis* and this clear limitation should be considered by assessing the prevalence of the other strain in the future studies (42, 54, 56). For the negative samples we did not rule out inhibition of PCR completely, which can make our results an underestimation of the true prevalence.

To conclude, the higher prevalence of human oral parasites in periodontal disease compared to healthy individuals appears to be more than a mere marker for the disease and might also be associated with disease severity and potential for progression. Thus, the dogmatic view of these oral parasites as commensals needs to be re-evaluated and their role cannot be neglected in light of the results of this study that supplement the recent articles that pointed to similar links. It is recommended to conduct future studies with the same molecular approach since the sole use of microscopy can lead to significant underestimation of the true prevalence of these oral parasites. Future studies are needed to assess the molecular epidemiology of these oral parasites and to test whether variations in strains that do exist, have a significant contribution in health and disease (61). Regional specificity for *T. tenax* appeared to be existent in Jordanian population and other studies in different geographic locations with different living standards might reveal if such specificity is genuine, or if it is only a spurious correlation involving an underlying factor.

## Materials and Methods

### Study design

The project was a prospective study with active enrolment of potential participants that took place during July 2019 to December 2019 at Jordan University Hospital (JUH). We sought to recruit study subjects from the following three categories: (1) Individuals with healthy gums (will be referred to as “healthy group” in the rest of manuscript), (2) Individuals with gingivitis (in this work, the term “gingivitis” will be applied to plaque-induced gingivitis, rather than non-dental-biofilm induced forms of gingivitis) and (3) individuals with periodontitis. The individuals with gingivitis and those with periodontitis (the disease group) were recruited from Periodontics Outpatient Clinics at JUH whereas healthy controls were recruited by active approach of the JUH staff that included dentists, laboratory technicians, nurses and students at the University of Jordan.

### Study participants

Each healthy individual was included in the study if the following criteria were met altogether: (1) Healthy gingiva on periodontal examination, (2) Bleeding index of less than 10%, and (3) No previous history of periodontal diseases. The criteria for the individuals with gingivitis and periodontitis were: (1) Diagnosis of the gum disease for the first time, and (2) No previous history of exposure to any kind of periodontal therapy (scaling or root planing). The presence of one of the following criteria resulted in non-inclusion of the potential participant in the study: (1) Pregnant woman, (2) Previous history of periodontal treatment, (3) Non-dental-biofilm induced forms of gingivitis, (4) Presence of dental implants, and (5) Orthodontics treatment.

### Paper-based questionnaire on possible risk factors for periodontal disease

Data from the study participants were collected using a paper-based questionnaire that comprised 17 different items (Supplementary File 1). The study participants’ data included: age, gender, body mass index (BMI), monthly income, and dental care level, history of smoking, alcohol consumption, diabetes mellitus, family history of gum disease and medications history. In addition, nine questions were used to assess the stress-related factors, with each positive response given a single point yielding a stress score that ranged from nil to nine.

### Ethical considerations

This study was approved by the School of Medicine and the School of Graduate Studies, University of Jordan. Ethical approval was obtained from the Institutional Review Board (IRB) at JUH (Ref. No. 239/2019). A written and signed informed consent was obtained from all individuals who agreed to participate in the study following full explanation of the study objectives and the procedure of obtaining the samples. In addition, the work was conducted according to the principles of good clinical practice that have their origin in the declaration of Helsinki and all individual data were treated with confidentiality.

### Classification of study subjects into healthy, gingivitis or periodontitis groups

The diagnosis of gingivitis and periodontitis was based on diagnostic guidelines that were set by the 2018 new classification scheme for periodontal and peri-implant diseases and conditions (11). The details are provided in (Supplementary File 2).

### Specimen collection and microscopic examination

Salivary and dental plaque specimens were obtained from each study subject. Supra-gingival plaque was removed then the sample was taken from the deepest periodontal pocket. Dental plaque samples were collected using the UNC periodontal probe (15 mm). For participants with furcation involvement, the sample was taken using Nabers probe from the furcation. For each participant, the salivary and dental plaque specimens were mixed together in a sterile tube. A single drop of the mixture was used for immediate wet mount examination using wide-field light microscopy, while the tubes were stored at −20°C for DNA extraction and amplification. For wet mount examination, the entire cover slip was inspected with 10× objective through a zigzag path to ensure scanning the sample entirely and the suspicious objects (based mainly the criteria defined by Bonner *et al*.) were examined at 40× objective (34).

### DNA extraction and amplification

DNA purification was done using QIAamp DNA Mini Kit (QIAGEN) according to manufacturer’s instructions. Briefly, the saliva/dental plaque specimens were brought to room temperature and mixed well, followed by adding 20 μL of proteinase k to a total of 200 μL of the specimen. If the specimen volume was less than 200 μL, we added a proper volume of phosphate buffered saline to reach a final volume of 200 μL. This was followed by adding 200 μL of buffer AL to the sample/proteinase k and vortexing for until a homogenous solution was formed. The mixture was then incubated at 56 °C for ten minutes. After that, 200 μL absolute ethanol was added to the lysed sample a mixed by vortexing for 15 seconds followed by its transfer into QIAamp Mini spin column. This was followed by centrifugation at 6000 × *g* for one minute. Washing steps followed using buffers AW1 and AW2 and DNA elution was done using 200 μL of buffer AE and centrifugation at 6000 × *g* for one minute.

For the detection of oral parasites, two sets of PCR primers were used. For *E. gingivalis*, we used the same set of primers utilized by Bonner *et al*. with minor modification of the reverse primer as follows: forward primer (5′-AGGAATGAACGGAACGTACA-3′) and reverse primer (5′-CCATTTCCTTCTTCTATTGTTTMAC-3′) with a product size of 203 bases(34). For *T. tenax*, we used the same set of primers utilized by Kikuta *et al*. as follows: PT3 forward primer (5′-AGTTCCATCGATGCCATTC-3′) and PT7 reverse primer (5′-GCATCTAAGGACTTAGACG −3′) with product size of 776 bases (52). The PCR mix comprised 5 μL of the DNA eluate, 5 μL of 5×FIREPol Master Mix (Solis BioDyne), 1 μL of each primer and 13 μL of DNase/RNase free water. The steps of PCR were as follows: Initial denaturation for 3.5 minutes at 94°C, 40 cycles of 1 minute at 94°C for denaturation, 1 minute at 60°C for primer annealing, 1 minute at 72°C for elongation, a final elongation step for 5 minutes at 72°C (34, 52, 62).

A volume of 6 µL of the final product was assessed using for 2% agarose gel electrophoresis for evaluation of the DNA product sizes. Proper positive and negative extraction and PCR controls were used to ensure the quality of DNA extraction and PCR and to rule out contamination. Positive controls were taken from periodontitis patients who were positive for *E. gingivalis* and *T. tenax* by microscopy and that yielded the correct band sizes, while the negative control was nuclease-free water. The housekeeping gene actin beta (*ACTB*) with accession number (NG_007992.1) was used to assess PCR inhibition of the sample and to ensure the efficiency of the DNA extraction procedure with the following primers: forward 5’ GTCCTGTGGCATCCACGAAA 3’ and reverse 5’ AGTGAGGACCCTGGATGTGAC 3’ and PCR product size of 265 bases.

### Statistical analysis

Data generated from the study were cleaned using Microsoft Excel and uploaded to IBM SPSS Statistics 22.0 for Windows. Chi-squared test (χ^2^ test), Mann-Whitney (M-W), Kruskall-Wallis (K-W) and linear-by-linear test for association (LBL) tests were used when appropriate and the statistical significance was considered for p<0.050. To analyse the patterns associated with higher likelihood of having periodontal disease as a whole and per disease state (gingivitis and periodontitis), we conducted multinomial logistic regression analysis using variables that were classified into dichotomous outcomes. Confidence intervals of percentages (95% CI) were calculated using modified Wald method through GraphPad calculator available freely online through the following link: https://www.graphpad.com/quickcalcs/ConfInterval1.cfm

## Data Availability

The datasets generated during and/or analysed during the current study are available from the corresponding author on reasonable request

## Acknowledgments

We would like to thank all the individuals who agreed to participate in the study. In addition, we would like to thank the Staff of the Department of Dentistry and the Department of Clinical Laboratories and Forensic Medicine at JUH for their help and support. Special thanks to Dr. Omar Alkaradsheh, Dr. Nicola Barghout, Dr. Mais Al-Ashqar and Dr. Belal Al-Azab for facilitating sample collection and laboratory work.

## Author Contributions

**Conceptualization:** AY, AM, GÖŞ, JSR and MS

**Funding acquisition:** AM

**Supervision:** AM, JSR and MS

**Data collection:** AY, DD and DT

**Sample collection:** AY, DD and DT

**Laboratory work:** AY

**Data analysis and interpretation:** All authors

**Statistical analysis:** MS

**Prepared tables:** MS

**Writing - original draft:** MS

**Writing - review & editing:** All authors

